# The impact of COVID-19 pandemic on influenza surveillance: a systematic review and meta-analysis

**DOI:** 10.1101/2022.03.31.22273236

**Authors:** Sasidharanpillai Sabeena, Nagaraja Ravishankar, Sudandiradas Robin

## Abstract

**Background:** Influenza activity was reported to be below the seasonal levels during the COVID-19 pandemic globally. However, during the SARS-CoV-2 outbreak, the routine real-time surveillance of influenza like illness (ILI) and acute respiratory infection (ARI) was adversely affected due to the changes in priorities, economic constraints, repurposing of hospitals for COVID care and closure of outpatient services.

**Methods:** A systematic review and meta-analysis were carried out to assess the pooled proportion of symptomatic cases tested for influenza virus before the current pandemic in 2019 and during the pandemic in 2020/21. An electronic search of PubMed/MEDLINE, Scopus and Google Scholar was carried out for the articles reporting the impact of the COVID-19 pandemic on Influenza surveillance among humans using search terms. The study was designed based on PRISMA guidelines and the meta-analysis was performed to synthesise the pooled proportion of patients sampled for influenza with 95% confidence interval (CI).

**Results:** The nine qualified studies from the WHO-European region, Canada, Japan, Germany, Italy, Spain, South Africa and the United States were pooled by random-effects meta-analysis. The overall pooled proportion of symptomatic cases sampled for influenza surveillance before and during the pandemic was 2.38% (95% CI 2.08%-2.67%) and 4.18% (95% CI 3.8%-4.52%) respectively. However, the pooled proportion of samples tested for influenza before the pandemic was 0.69% (95% CI 0.45-0.92%) and during the pandemic was 0.48% (95% CI 0.28-0.68%) when studies from Canada were excluded.

**Conclusion:** The meta-analysis concludes that globally there was a decline in influenza surveillance during the COVID-19 pandemic except in Canada.

**Key Messages:**

- The nine observational studies from Europe, Canada, Japan, South Africa and the United States were qualified for the meta-analysis
- A steep decline in the seasonal influenza activity in both northern and southern hemispheres was observed
- Almost double the number of symptomatic cases were sampled as part of influenza surveillance during the current pandemic in Canada
- Except in Canada, a decline in influenza surveillance globally during the COVID-19 pandemic was observed

## Introduction

Severe acute respiratory syndrome corona virus-2 (SARS-CoV-2) virus is genetically linked to deadly SARS coronavirus, which emerged in 2002 and disappeared within eighteen months. SARS-CoV-2 and influenza viruses affecting the respiratory systems have the same transmission route, and various control measures to combat the current pandemic mitigate the spread of other respiratory viruses such as influenza and respiratory syncytial virus.^1^The diagnosis of influenza cases was 99% lower in 2020 than in the previous influenza seasons.^2,3^

Influenza is a systematically monitored viral disease due to the ongoing threat of epidemics and pandemics. A network of laboratories known as Global Influenza Surveillance and Response System (GISRS) was established in 1952 to update the circulating influenza strain variation information. This system facilitates the prompt identification and implementation of preventive measures against impending influenza activity in the community. The changing priorities in infectious disease surveillance and resource constraints during the Coronavirus disease-2019 (COVID-19) pandemic have negatively impacted the surveillance. Many countries have witnessed a short-term interruption or delayed influenza data reporting during the COVID times.

Meanwhile, a study from Canada reported a drastic fall in the number of other seasonal non-SARS-CoV-2 respiratory viral infections, including influenza and respiratory syncytial virus (RSV).^4^The surveillance data from the United States and Australia reported low influenza activity during 2020/2021.^5,6^ Surveillance is mandatory to identify and respond to early Influenza outbreaks of pandemic potential, and sentinel surveillance among a defined population provides data with the highest quality^7^.

This systematic review and meta-analysis aimed to evaluate the pooled proportion of patients sampled for influenza before and during the COVID-19 pandemic. This quantitative meta-analysis emphasises the importance of maintaining influenza surveillance during challenging times.

## 2 Methods

This systematic review and meta-analysis were started after excluding registered or ongoing systematic reviews regarding the impact of the COVID-19 pandemic on influenza surveillance in the PROSPERO database.

The study protocol was registered in PROSPERO database (CRD42021296702) and can be accessed at www.crd.york.ac.uk/PROSPERO/display_record.asp?ID=CRD42021296702. An electronic search of PubMed/MEDLINE, Scopus and Google Scholar was carried out for the articles in English concerning the impact of the COVID-19 pandemic on Influenza surveillance among humans between January 2019 and December 2021. The study protocol was designed based on Preferred Reporting Items for Systematic Reviews and Meta-analyses (PRISMA) guidelines updated in May 2020.^8^ The meta-analysis component was modified appropriately to synthesise the pooled proportion of patients sampled for influenza surveillance before the COVID-19 pandemic in 2019 and during the pandemic among the catchment population.

### 2.1 Description of the condition

#### Influenza-like illness (ILI)

A case is defined as ILI if the symptoms begin within the past ten days with a measured fever of 38°C or more, cough (respiratory infection).^9^

#### Acute Respiratory Infection (ARI)

Some countries use ARI instead of ILI for surveillance of respiratory viruses, including influenza in humans.

#### SARI

A case is defined as SARI if the symptoms begin within the past ten days with fever (measured≥ 38?) and respiratory infection (cough) necessitating hospitalization^9^.

### 2.2 Study Protocol

An electronic search of PubMed/MEDLINE, Scopus and Google Scholar was carried out for all the articles published between January 2020 and December 2021 concerning the impact of the COVID-19 pandemic on Influenza surveillance. The relevant articles in English involving human subjects were identified using search terms such as “impact” AND “COVID-19” OR “SARS-CoV-2” AND “influenza surveillance” NOT “vaccination”.

### 2.3 Inclusion process and criteria

Original research articles published in peer-reviewed scientific journals reporting the number of ILI/ARI/SARI cases sampled for influenza among the catchment population, as part of influenza surveillance at the sentinel, non-sentinel and diagnostic surveillance units before the COVID-19 pandemic in 2019 and during the pandemic were included. Observational studies such as cross-sectional and cohort studies reporting the number of patients screened for influenza among the target population were included. The studies reporting only the influenza positivity were excluded.

### 2.4 Data extraction

A validated proforma including the first author, year of publication, region, study design, number of ILI/ARI/SARI cases attending the influenza surveillance centres before the pandemic in 2019, symptomatic cases during the pandemic 2020/2021 and the target population of the study area was prepared. A three-stage selection process was carried out for the final inclusion of the studies. One reviewer assessed the titles from the records for the relevance for inclusion in the study (n=5384). Studies applicable to the review were moved to the second stage after excluding irrelevant topics and duplicates (n=155). In the second stage, the abstracts of the studies were obtained and were independently analysed by two reviewers (n=94). After reviewing the abstracts, full texts of studies were retrieved, which were scrutinised by two reviewers independently (n=55). The corresponding authors were contacted electronically if further interpretation was needed. Manual library searches for articles in peer-reviewed journals were carried out, and references of retrieved articles were reviewed to increase the search sensitivity. The study selection process was depicted in the PRISMA chart (**Figure 1**). The last date of the search was on January 31, 2022.

**Figure 1:**
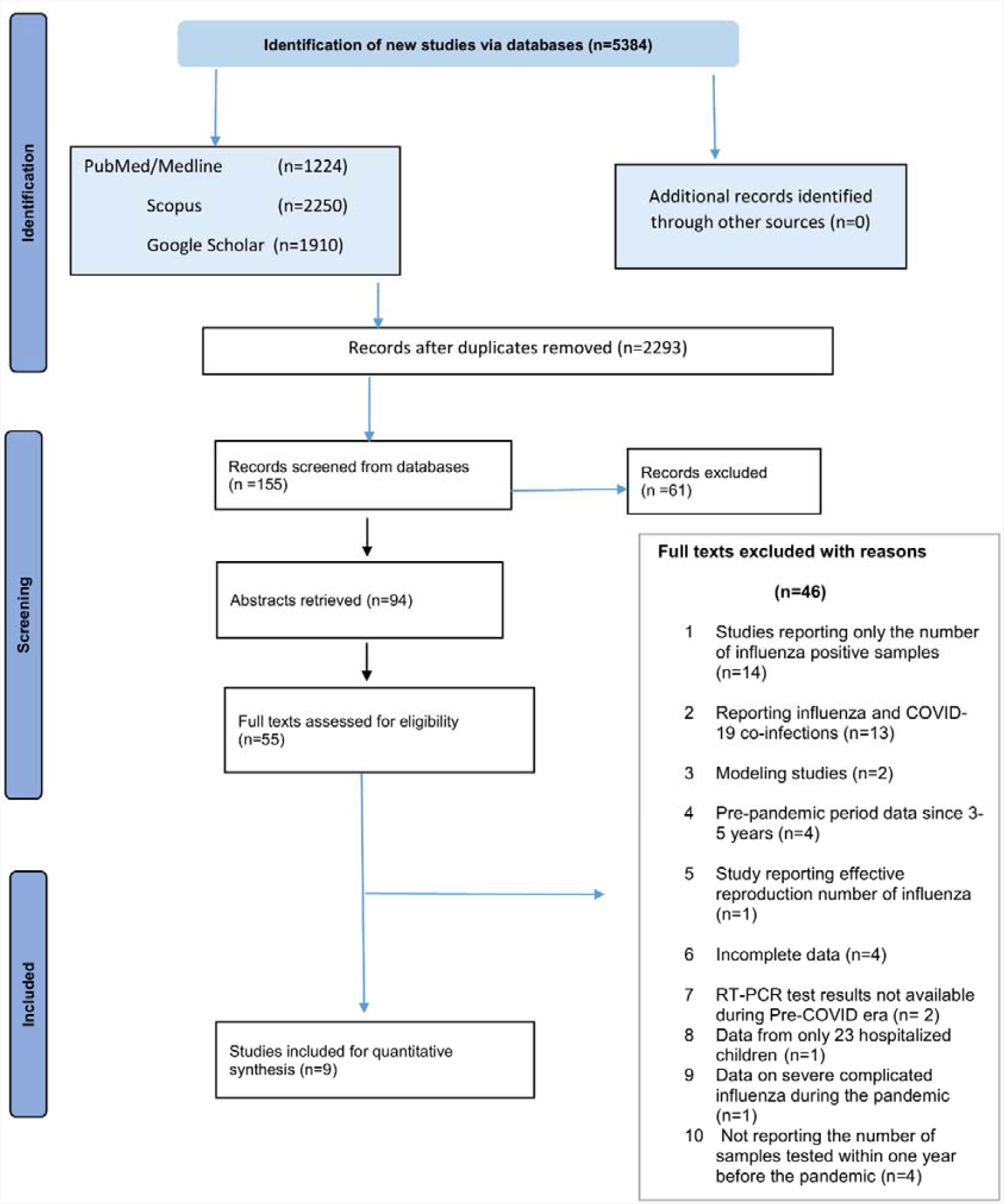
PRISMA 2020 flow diagram for updated systematic reviews which included searches of databases, registers and other sources. The flow diagram illustrates the number of studies identified, screened, abstracts/full-text articles included/excluded for the systematic review and meta-analysis.

### 2.5 Risk of bias (quality) assessment in individual studies

To assess the risk of bias in individual studies (quality assessment), chosen after the abstract and content review, the National Institutes of Health checklist for observational, cohort and cross-sectional studies was used^10^. The studies with a minimum score of eight or above, seven, or five or less than five “Yes responses” were considered good, fair, and poor quality, respectively. For cross-sectional studies, question numbers 1, 2, 3, 4, 5 and 11 were applicable. The responses to the remaining eight questions (6-10, 12, 13, 14) were marked as not applicable (NA). The studies with six “Yes” responses were considered good, and those with four /five were taken as fair. The studies with less than four “Yes responses” were considered poor quality. The quality of the studies was assessed independently by two reviewers.

### 2.6 Statistical analysis

The meta-analysis was accomplished in STATA version 13.0 (College Station, Texas 77, 845 USA). The forest plots were constructed using metaprop package in STATA. A substantial amount of heterogeneity across the studies was expected as the included studies were observational. The pooled proportion of symptomatic cases of influenza-like illness, acute respiratory illness or severe acute respiratory illness presented to surveillance centres out of the catchment population before the pandemic in 2019 and during the pandemic was reported with 95% CI along with Chi-square statistic (Q statistic) and I^2^ index to quantify the heterogeneity. The I^2^ value ranging between 0% to 24% specifies consistency. I^2^ values of 25%-49% imply low heterogeneity, and 50-74% point toward moderate heterogeneity. The I^2^ value varying between 75%-100% is indicative of high heterogeneity.^11^

### 2.7 Assessment of Publication bias

The egger’s test was used to analyse the publication bias. Weighted linear regression with standardised effect estimate and precision were considered the dependent and independent variable, respectively. In the present study, the log_e_ proportion of patients under surveillance and precision were considered the effect estimate and 1/standard error of log_e_ proportion rate, respectively. Weights were allotted using the inverse variance approach (1/variance of the effect estimate). A statistically significant bias coefficient is the evidence for publication bias.

## Results

### 3.1 Included studies

We identified and screened 155 articles from the databases including the abstracts of 94 articles. Out of these, full texts of 55 articles were retrieved and systematically reviewed. The nine qualified studies for the meta-analysis were from the WHO-European region, Canada, Japan, Germany, Italy, Spain, South Africa and the United States, comprising 448,423 symptomatic cases sampled before the pandemic in 2019 and 1,270,518 cases sampled during the pandemic in 2020-2021^2,7,12–18^. Except for one study from Spain, which did not specify the months during which cases were sampled, all the included studies were qualified as good.^16^ As per the US-CDC influenza surveillance data between 2009 and 2019, approximately 8-9% of the US population is under routine influenza sentinel-surveillance.^19^ Meanwhile, about 1% of the total population in Germany and 2.5% of the total population in Spain are under sentinel influenza surveillance in 2019.^20^ The comprehensive surveillance data from the WHO-European region was reported by Adlhoch et al.^7^ The cases with influenza illness under the influenza sentinel surveillance by the representative national network of primary care practitioners is highly variable in WHO European region^20^. If the studies lack information on the catchment population, 6% of the region’s total population was taken as the denominator as most of the European countries cover around 6% population in sentinel surveillance.^7,12,15,17,18^ The qualified study from Hokkaido Prefecture in Japan carried out non-sentinel surveillance of respiratory infection among hospitalised children below 15 years. The catchment population of children aged less than 15 was 11.4% of the total population^13^. Amongst the qualified studies, the number of cases under surveillance varied between 2128 and 508,533 before the pandemic in 2019. As in **Table 1** more symptomatic cases were sampled in WHO-European region and Canada in 2019. Canada tested the highest number of symptomatic cases for influenza during the pandemic. Compared to the pre-pandemic period, the two qualified studies from Canada tested more than double the number of ILI cases for influenza as part of sentinel hospital and laboratory-based studies^12,18^. Park et al. from Canada reported the proportion of cases under influenza surveillance in the whole of 2019 (before pandemic) and the entire 2020.^12^ Meanwhile, another study from Canada reported the sentinel laboratory surveillance data during the three corresponding months in 2019 and 2020.^18^

**Table 1:**
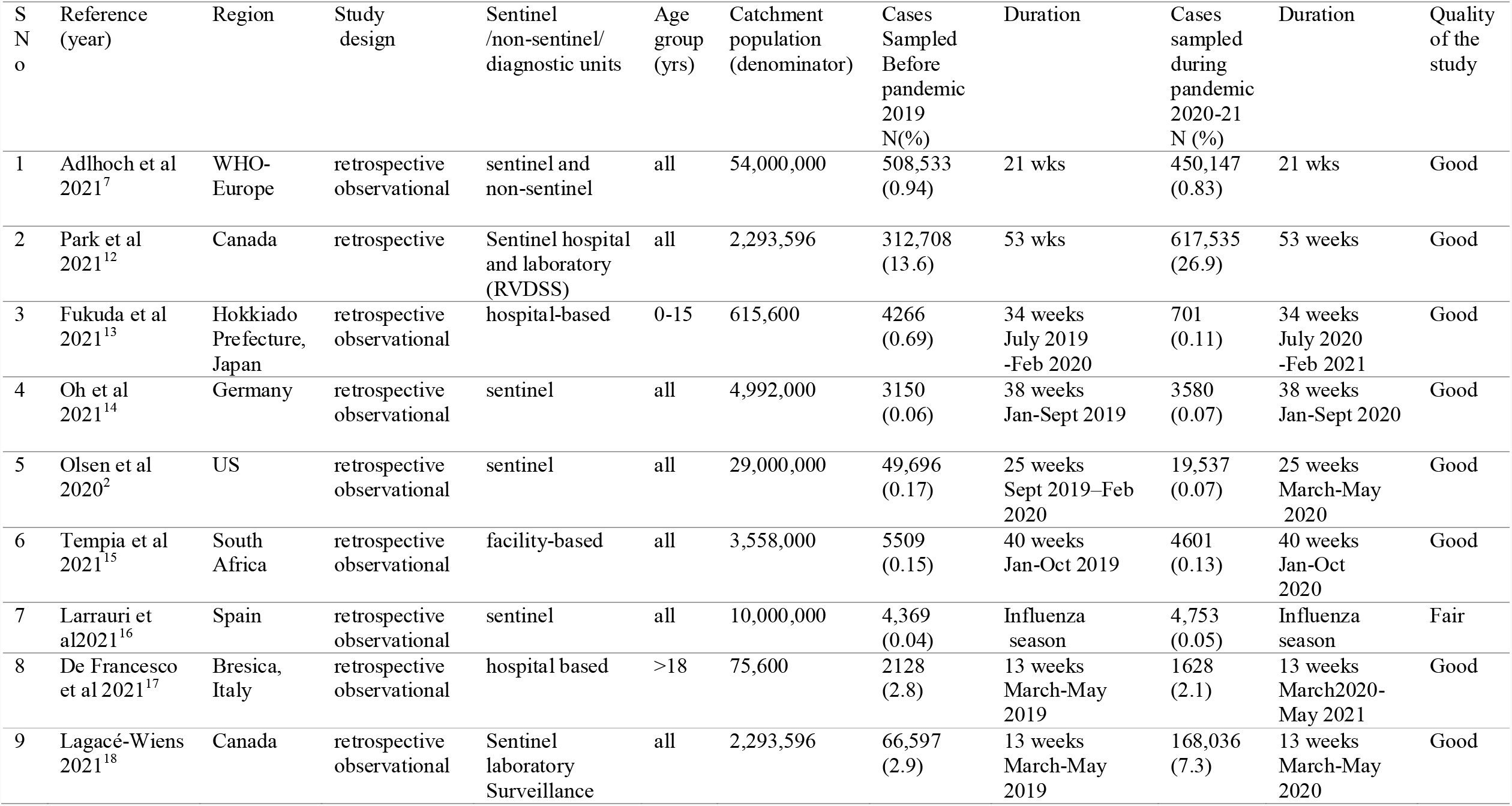
The characteristics of the studies included for the meta-analysis.

| S<br>N<br>o | Reference<br>(year) | Region | Study<br>design | Sentinel<br>/non-sentinel/<br>diagnostic units | Age<br>group<br>(yrs) | Catchment<br>population<br>(denominator) | Cases<br>Sampled<br>Before<br>pandemic<br>2019<br>N(%) | Duration | Cases<br>sampled<br>during<br>pandemi<br>c<br>2020-21<br>N (%) | Duration | Quality<br>of the<br>study |
| --- | --- | --- | --- | --- | --- | --- | --- | --- | --- | --- | --- |
| 1 | Adlhoch et al<br>2021<sup>7</sup> | WHO-Europe | retrospective<br>observational | sentinel and<br>non-sentinel | all | 54,000,000 | 508,533<br>(0.94) | 21 wks | 450,147<br>(0.83) | 21 wks | Good |
| 2 | Park et al<br>2021<sup>12</sup> | Canada | retrospective | Sentinel<br>hospital and<br>laboratory<br>(RVDSS) | all | 2,293,596 | 312,708<br>(13.6) | 53 wks | 617,535<br>(26.9) | 53 weeks | Good |
| 3 | Fukuda et al<br>2021<sup>13</sup> | Hokkaido<br>Prefecture,<br>Japan | retrospective<br>observational | hospital-based | 0-15 | 615,600 | 4266<br>(0.69) | 34 weeks<br>July 2019<br>-Feb 2020 | 701<br>(0.11) | 34 weeks<br>July 2020<br>-Feb 2021 | Good |
| 4 | Oh et al<br>2021<sup>14</sup> | Germany | retrospective<br>observational | sentinel | all | 4,992,000 | 3150<br>(0.06) | 38 weeks<br>Jan-Sept 2019 | 3580<br>(0.07) | 38 weeks<br>Jan-Sept 2020 | Good |
| 5 | Olsen et al<br>2020<sup>2</sup> | US | retrospective<br>observational | sentinel | all | 29,000,000 | 49,696<br>(0.17) | 25 weeks<br>Sept 2019–Feb<br>2020 | 19,537<br>(0.07) | 25 weeks<br>March-May<br>2020 | Good |
| 6 | Tempia et al<br>2021<sup>15</sup> | South<br>Africa | retrospective<br>observational | facility-based | all | 3,558,000 | 5509<br>(0.15) | 40 weeks<br>Jan-Oct 2019 | 4601<br>(0.13) | 40 weeks<br>Jan-Oct<br>2020 | Good |
| 7 | Larrauri et<br>al 2021<sup>16</sup> | Spain | retrospective<br>observational | sentinel | all | 10,000,000 | 4,369<br>(0.04) | Influenza<br>season | 4,753<br>(0.05) | Influenza<br>season | Fair |
| 8 | De Francesco<br>et al 2021<sup>17</sup> | Brescia,<br>Italy | retrospective<br>observational | hospital based | >18 | 75,600 | 2128<br>(2.8) | 13 weeks<br>March-May<br>2019 | 1628<br>(2.1) | 13 weeks<br>March 2020-May 2021 | Good |
| 9 | Lagacé-Wiens<br>2021<sup>18</sup> | Canada | retrospective<br>observational | Sentinel<br>laboratory<br>Surveillance | all | 2,293,596 | 66,597<br>(2.9) | 13 weeks<br>March-May<br>2019 | 168,036<br>(7.3) | 13 weeks<br>March-May<br>2020 | Good |

Our meta-analysis included data from sentinel, non-sentinel and hospital-based surveillance centres in the context of an ongoing pandemic. The number of symptomatic cases sampled from hospitals was less than sentinel surveillance samples. Three studies enrolled cases as part of non-sentinel surveillance.^13,15,17^ Even though all age groups were included in seven qualified studies, there was a discrepancy in the age groups enrolled for the surveillance.

### 3.2 Meta-analysis

The overall pooled proportion of symptomatic cases undergone influenza testing before the pandemic in 2019 was 2.38% (95% CI 2.08%-2.67%) as in **figure 2**. The **figure 3** depicts the pooled proportion of cases sampled during the pandemic in 2020/2021, which was 4.18% (95% CI 3.84%-4.52%). However, the pooled proportions of samples tested before the pandemic in 2019 and during the pandemic were 0.69% (CI 0.45%-0.92%) and 0.48% (CI 0.28%-0.68%), respectively, when the studies from Canada were excluded (**figure 4 & figure 5)**. As the I^2-^value was>90%, high heterogeneity was observed between the studies and random-effects model was used for pooling.

**Figure 2:**
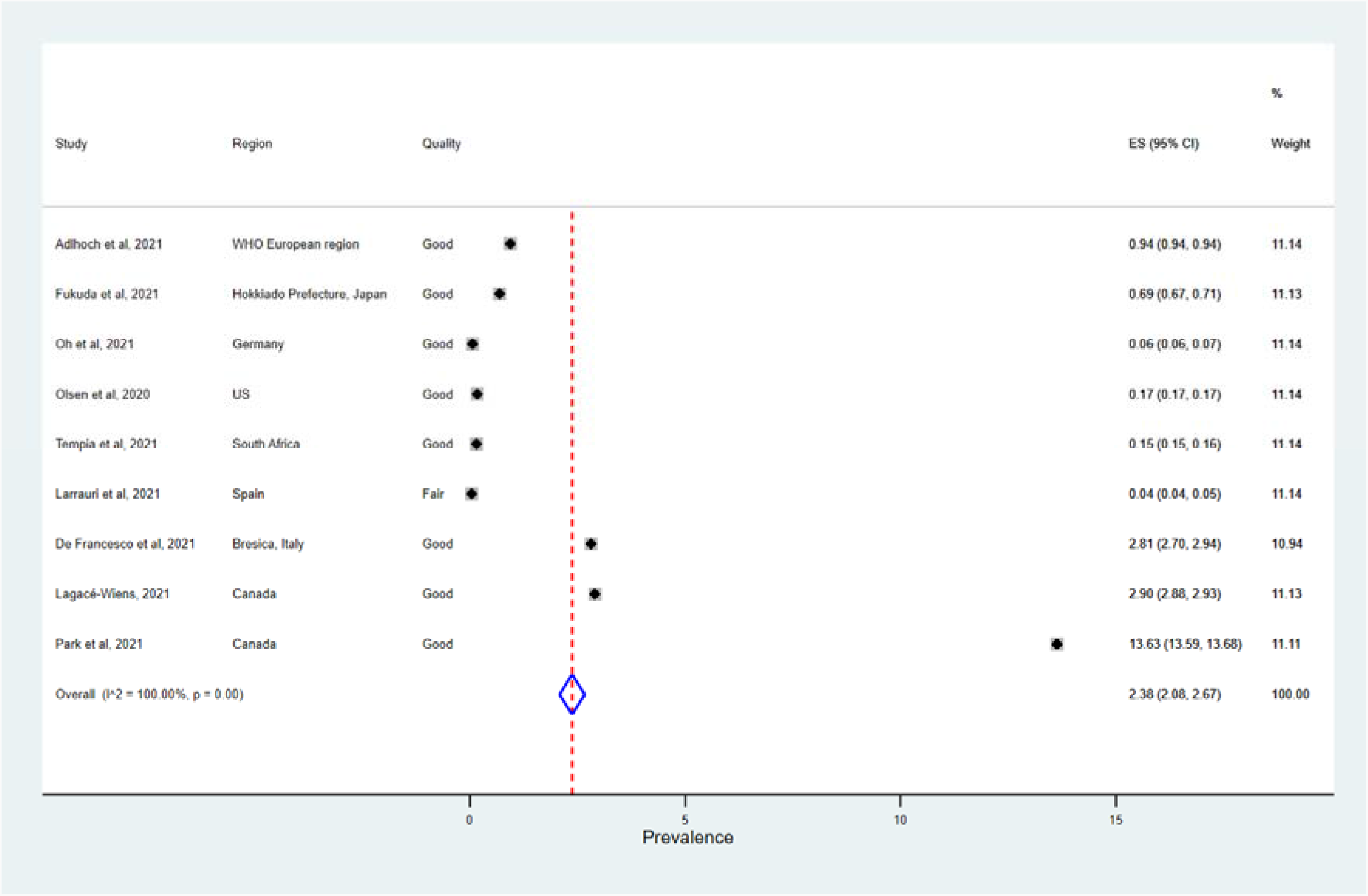
The Forest Plot of the summary effect size (Proportion of symptomatic cases tested for influenza among the catchment population before the COVID-19 pandemic) using random effects model amongst the target population. Squares indicate the effect size of individual studies and the extended lines denote 95% confidence intervals (CI). Sizes of squares imply the weight of studies based on sample size using a random effects analysis. The diamond data indicates pooled prevalence. Test of heterogeneity: I^2=^100%, *p*-value=0.00

**Fig 3.**
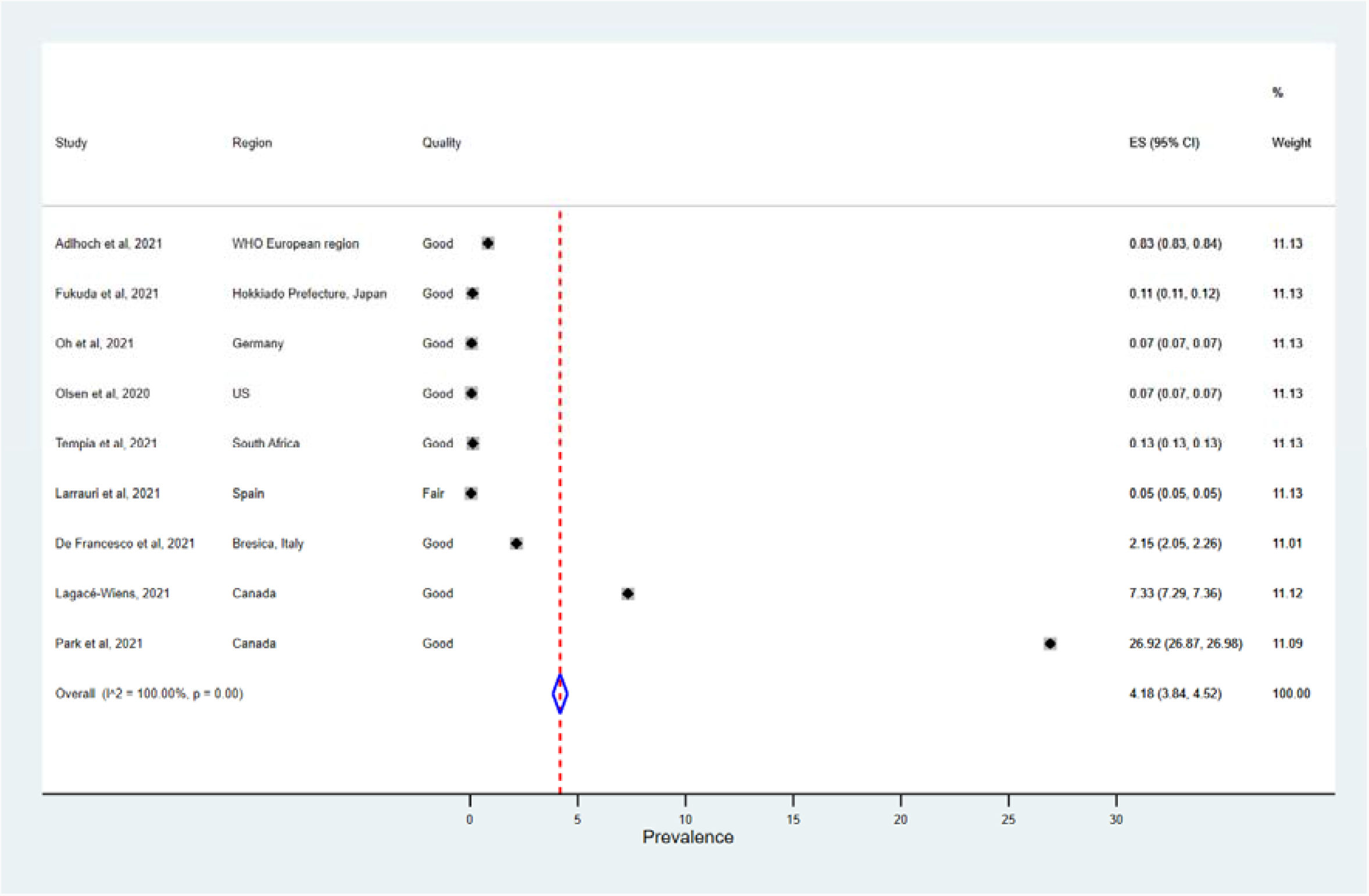
The Forest Plot of the summary effect size (Proportion of symptomatic cases screened for influenza among the catchment population during the COVID-19 pandemic) using random effects model. Squares indicate the effect size of individual studies and the extended lines denote 95% confidence intervals (CI). Sizes of squares imply the weight of studies based on sample size using a random effects analysis. The diamond data indicates pooled prevalence. Test of heterogeneity: I^2=^100 %, *p*=0.00.

**Figure 4:**
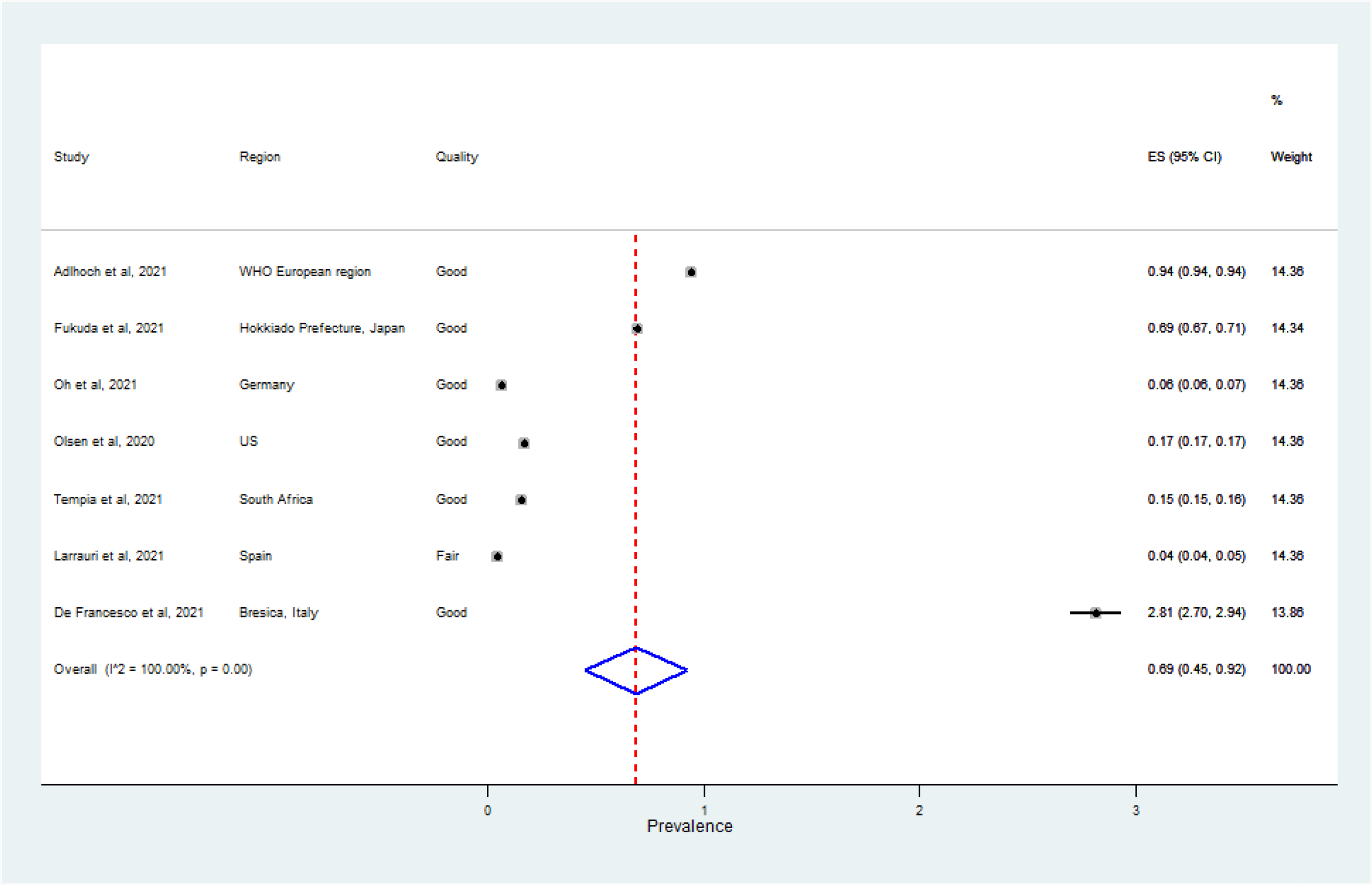
The Forest Plot depicting the summary effect size (Proportion of symptomatic cases tested for influenza among the catchment population before the COVID-19 pandemic) using random effects model amongst the target population after eliminating the studies from Canada. Test of heterogeneity: I^2=^100.00%, *p*-value=0.00

**Figure 5:**
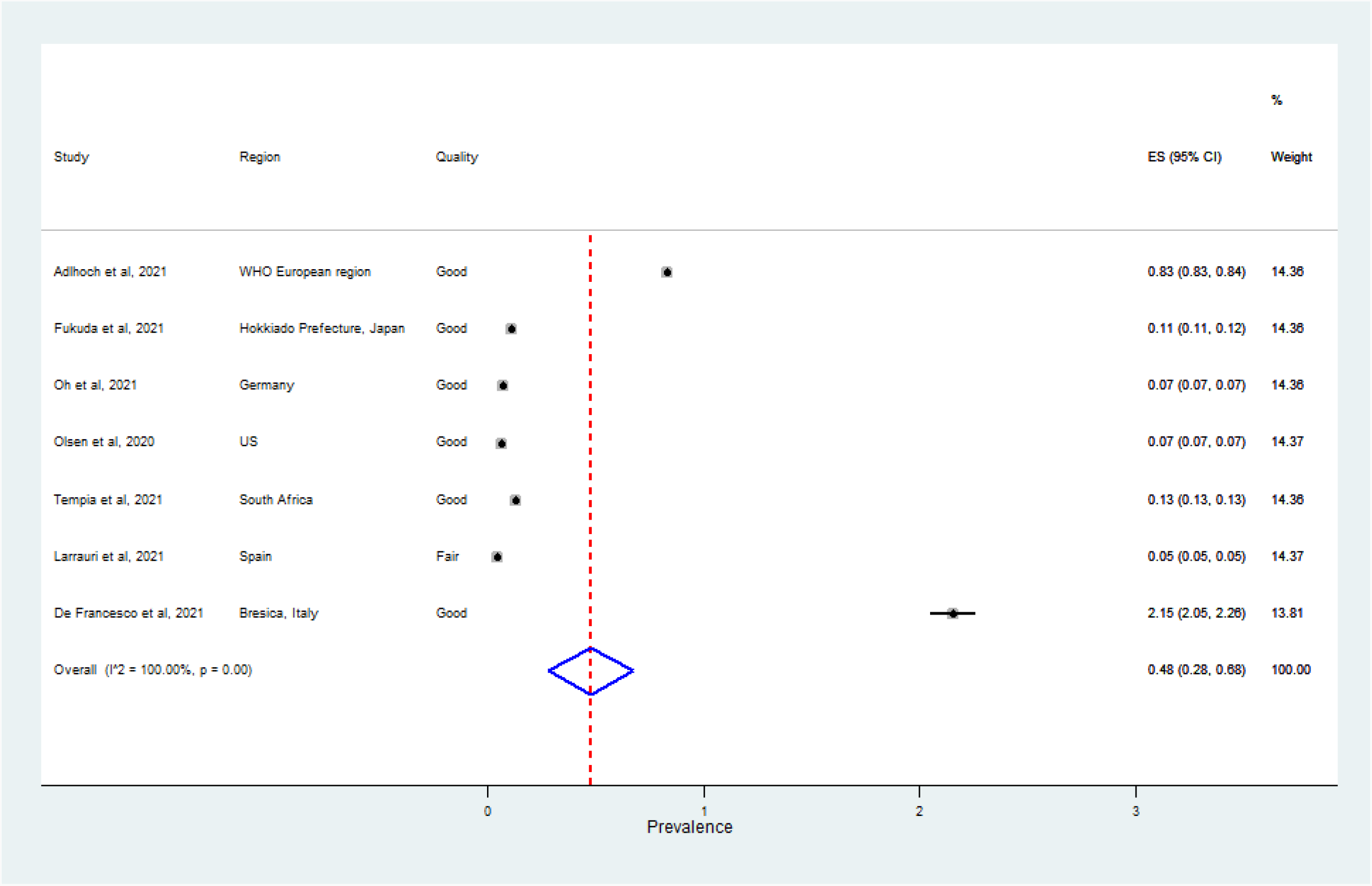
The Forest Plot depicting the summary effect size (Proportion of symptomatic cases tested for influenza among the catchment population during the COVID-19 pandemic) using random effects model amongst the target population after eliminating the studies from Canada. Test of heterogeneity: I^2=^100.00%, *p*-value=0.00

### 3.3 Publication bias

There was no publication bias as the *p*-value for the bias coefficient was not statistically significant as shown Supplementary Table 1 (**Table 2**).

**Table 2:**
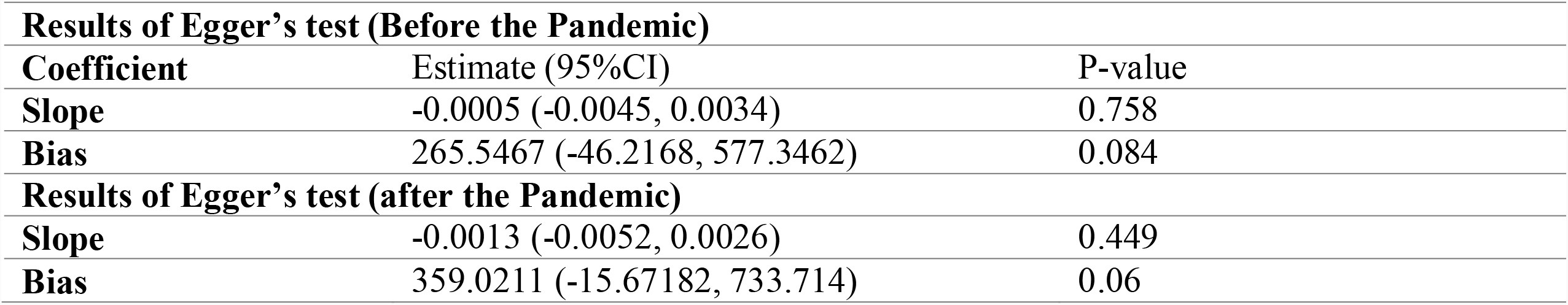
The table depicts the results of Egger’s test for publication bias.

| **Results of Egger's test (Before the Pandemic)** |  |  |
| --- | --- | --- |
| **Coefficient** | **Estimate (95%CI)** | **P-value** |
| **Slope** | -0.0005 (-0.0045, 0.0034) | 0.758 |
| **Bias** | 265.5467 (-46.2168, 577.3462) | 0.084 |
| **Results of Egger's test (after the Pandemic)** |  |  |
| **Slope** | -0.0013 (-0.0052, 0.0026) | 0.449 |
| **Bias** | 359.0211 (-15.67182, 733.714) | 0.06 |

## 4 Discussion

Globally there was a decline in influenza surveillance during the COVID-19 pandemic except in Canada. Almost double the number of symptomatic cases were sampled as part of influenza surveillance during the current pandemic in Canada^12,18^ compared to the pre-pandemic period. This might be attributed to dual testing strategy for influenza and SARS-CoV -2 as part of capacity building during the current pandemic.^21^ The WHO European region testing a large number of influenza samples reported a decline in surveillance samples during the current pandemic. This region noted the lowest number of influenza types and subtypes since the establishment of GISRS in 1952.^7^ In the US, the surveillance for influenza and the influenza test positivity rate declined by 61% and 98% respectively.^2^Similarly, the seasonal influenza positivity declined by 64% in China ^22^ and the sentinel surveillance of influenza was interrupted in Italy^23^. Australia witnessed consistently low influenza-like illness activity (ILI) since July 2021^24^. However, upgrading the established influenza surveillance system empowered the European countries for the monitoring of SARS-Co-V-2.^25^ As per WHO, influenza surveillance and SARS-CoV-2 monitoring have to be carried out along with syndromic surveillance of SARI cases hospitalised during the pandemic. Case definitions specific to COVID-19/ARI/ILI/SARI have to be incorporated into the case reporting and sample submission forms.^26^ A subset of SARS-CoV-2 samples have to be tested for influenza.

Countries in the northern hemisphere reported a fall in influenza cases during the influenza season spanning between October and April in 2020.^22,27–29^ During the regular influenza season in the southern hemisphere from April to September Australia, New Zealand, South Africa, and Chile also observed low influenza activity.^15,30,31^ Upon systematic review, the studies from China, Taiwan, Korea, Singapore, Japan and Bangladesh reported low influenza activity during the COVID-19 pandemic.^27,29,32–35^Meagre test positivity rates for all respiratory viruses, including influenza and respiratory syncytial virus (RSV), were reported from Canada.^4^ A recent study from Canada reported no influenza-related hospital admissions or deaths among paediatric cases aged 16 years and below during the 2020/21 influenza season.^36^ China witnessed the lowest test positivity rates of respiratory viruses over the past decade during the current pandemic.^37^ The test positivity was consistently low for influenza and human metapneumovirus with increased detection rates for rhinovirus, coronavirus, adenovirus, RSV, human parainfluenza virus, and Boca virus when the non-pharmaceutical interventions (NPI) were relaxed in China. There was a substantial reduction in detecting all types and subtypes of influenza in the WHO-European region during the 2020/2021 influenza season.^38^ Due to the execution of strict measures to control the COVID-19, low influenza and respiratory syncytial virus (RSV) activity in South Africa was observed. Even though there was an increase in the detection rate of RSV with the easing of public health measures, the influenza positivity remained at very low levels.^15^However, an increase in the circulation of rhinovirus was observed in Canada, as in Japan and Turkey^39,40^ which is often attributed to the greater environmental stability of non-enveloped viruses and greater efficacy of face masks in filtering out enveloped viruses.^41^

There was a significant disparity in the number of weeks during which the samples were procured for surveillance. Some studies included data from influenza season and few retrieved data from late winter. The data were based on sentinel, non-sentinel and hospital-based surveillance in the context of an ongoing pandemic. Even though all the age groups were included in seven qualified studies, there was a discrepancy in the age groups enrolled for the surveillance. Five of the nine qualified studies included data from surveillance centres with a defined denominator. Meanwhile, there were no well-defined catchment population for hospital or facility-based surveillance. The number of symptomatic cases sampled from hospitals was less than that of sentinel surveillance samples.

Various public health measures such as hand sanitisation, face masking, social distancing, travel restrictions, working from home, school closures and changes in health-seeking behaviours contribute to in low influenza activity. During the early months of the pandemic, outpatient services and elective procedures were deferred worldwide in hospitals. Even after the relaxation of lockdowns, fewer patients with acute respiratory infections attended the hospitals mainly due to the fear of getting infected with SARS-CoV-2 and transmitting it to elderly family members. The changed priorities in healthcare seeking behaviours and economic constraints were also translated as reduced influenza surveillance, lower testing of samples for influenza and low positivity rates globally. Even sentinel physicians were reassigned for COVID duties contributing to lower influenza surveillance in many European countries such as Slovenia and Germany^7,14^.

A steep drop in the number of influenza cases during the current pandemic was attributed to non-pharmaceutical public health measures, modified health care seeking behaviours and testing priorities. However, reduced population exposure might result in low immunity to the influenza virus resulting in a rebound activity in the coming seasons.

Even during challenging times, sustained surveillance of influenza-like illness is essential to evaluate the geographical extent and sudden changes in circulating influenza strains globally. Optimisation of vaccines to circulating influenza strains is possible only by ensuring consistent influenza surveillance of the catchment population. The sparse data on the genetic and antigenic characterisation during 2020/2021 results in insufficient information on the vaccine composition for the coming influenza seasons. An innovative, cost-effective and sustainable strategy is needed to concentrate on influenza surveillance at the human-animal interface as many emerging and re-emerging infections in humans, including 2009 Pandemic influenza (H1N1 pdm09 virus), Nipah virus, Middle East Respiratory Syndrome Coronavirus (MERS Co-V) and SARS-CoV-2 are zoonotic. Data from four continents namely Africa, America, Asia, and Europe were included. The catchment population was well defined.

Even though we limited the review to articles in English, very few studies were excluded for that reason. Our systematic review observed a lack of data from the Middle East, South East Asia, Western Pacific and Latin American regions regarding the number of acute respiratory cases sampled for influenza surveillance before the COVID-19 pandemic in 2019. The studies from South East Asia and South America were omitted as the data regarding the number of samples tested in 2019 could not be retrieved, even after contacting the corresponding authors. Another limitation was the inaccuracy in the catchment population for hospital-based and non-sentinel surveillances. Due to a lack of studies enrolling all age groups from Japan, one hospital-based study among paediatric cases had to be included for the quantitative synthesis.

Globally there was a decline in influenza surveillance during the COVID-19 pandemic except in Canada. A steep decline in the seasonal influenza activity in both northern and southern hemispheres was observed. There is a need to have resilient ILI/SARI surveillance even during pandemics like COVID19 to recognise outbreaks by novel respiratory pathogens of pandemic potential.

## Data Availability

All data produced in the present study are available upon reasonable request to the corresponding author (Dr Sasidharanpillai Sabeena)

## Notes

### Competing Interest Statement

The authors have declared no competing interest.

### Clinical Protocols

http://www.crd.york.ac.uk/PROSPERO/display_record.asp?ID=CRD42021296702

### Funding Statement

This study did not receive any funding

### Summary of Updates

Senteces were edited in the results and Discusion parts

